# More Digital, More Diverse? How Digital Health Technologies Contribute to Representativeness of Clinical Trials

**DOI:** 10.1101/2025.07.28.25332315

**Authors:** Anatol-Fiete Näher, Linea Schmidt, Marvin Kopka, Matthias Schulte-Althoff, Akhyar Ahmed

## Abstract

The rapid adoption of digital health technologies (DHT) in clinical trials promises to enhance trial representativeness by eliminating geographical barriers and reducing participant burden. However, their actual impact on participant representation remains poorly understood. Our analysis of 68,206 US-based trials from ClinicalTrials.gov uncovers gaps in participant representation in trials utilizing DHT for data collection (DHTcd). While DHTcd trials achieved overrepresentation of female participants (+1.7% versus US Census data and +5.5% vs. non-DHT trials) and reduced underrepresentation of Black participants by 4.2% (vs. non-DHT trials), they showed underrepresentation of adults aged 65 years and older (−8.8% vs. non-DHT trials). These effects vary by therapeutic area. We furthermore find that DHTcd trials report group-specific sample compositions less frequently than non-DHT trials, suggesting that growing DHTcd adoption has not been systematically used to improve trial representativeness. We conclude that evidence-based and indication-specific implementation strategies are needed to enhance standards for DHT-based data collection in clinical trials and to prevent a widening digital divide in trial participation.

## Introduction

Technologies utilizing connectivity, computing platforms, software and sensors hold great potential for improving clinical trials. When used in healthcare, these technologies, such as smartwatches, fitness trackers, or mobile applications, are commonly referred to as ‘Digital Health Technologies’ (DHT) by the US Food and Drug Administration (FDA).^1^ The use of DHT in clinical trials has potential benefits in a number of areas: in addition to therapeutic and diagnostic applications, a key advantage of DHT is that data can be captured in interoperable formats and transferred seamlessly.^2,3^ This can possibly reduce the time needed for evidence generation and significantly increase the efficiency of clinical trials.^4^ Another distinctive advantage of DHT is that, unlike conventional data acquisition methods, data can be collected continuously, on a larger scale, and independent of a patient’s location.^4,5^ Thereby, DHT-based data acquisition methods (also called DHT collecting data, henceforth ‘DHTcd’) facilitate the generation of new biomedical knowledge. Such knowledge can be used, for instance, to develop digital biomarkers or to more accurately determine the desired and undesired effects of novel therapeutics or medical devices.^6^ The full benefits of using DHTcd to collect data in clinical trials can only be realised if DHTcd is used for all patients in interventional trials and observational studies. Accordingly, we define DHTcd trials as those in which DHT were used to capture endpoint data for every arm in interventional trials or data of interest from every patient included in observational studies.

A growing body of literature suggests that the use of DHTcd may also help improve representativeness of clinical trials.^7,8,5^ In the majority of available conventional trials, specific patient populations are not adequately represented or only represented in case numbers that do not permit subgroup-specific assessments of therapeutics or medical devices.^9,10^ For example, in the US population, women are overrepresented in US vaccine clinical trials, while patients over the age of 65 are underrepresented in these trials.^11^ The same is true for the representation of ethnic groups; for example, Black patients are underrepresented in US oncology trials, as measured by group-specific cancer incidence rates.^12^ These findings are essential for healthcare because not all therapeutic agents or medical devices have the same effect on all patient groups.^13–16^ Therefore, improving representation in clinical trials is crucial for developing targeted therapies by enabling the evaluation of a given therapeutic’s safety profile and mechanism of action across diverse patient groups.

A key reason why DHTcds can enhance trial representativeness and contribute to more targeted therapies is that their use allows trials to be decentralized. In many cases, geographical barriers constrain participation in clinical trials.^5^ For already underrepresented patient groups, such as the elderly population, travelling to study centers is often burdensome.^17^ Also women are often underrepresented in clinical trials and are more likely than men to be involved in childcare and to work in lower-paid jobs, making traveling to trial centers often too time-consuming and costly.^18,19^ In principle, decentralized trials with DHTcds have the potential to recruit underrepresented patient groups more successfully because the time and effort involved in travelling to trial centers is eliminated.^20,21^ In addition to decentralization, DHT can be used to implement digital interventions to build trust. Factors that have historically contributed to low trust in clinical research include misunderstanding of informed consent or actual trial processes and workflows, such as randomization into treatment and control arms.^3,17^ Novel digital formats, such as video consultations, for instance, can be used to offer help and information better tailored to the individual needs of trial participants, thereby increasing trust in clinical research processes and improving recruitment of previously disenfranchised patient populations.^22,23^ As recent experimental evidence suggests, trials with adequate representation also increase confidence in effective new treatments.^24^ A history of research misuse and restrictions on equitable access to healthcare services has led to disadvantaged populations, such as Black Americans, having limited trust in the healthcare system. This lack of trust has been linked to lower healthcare utilization and higher mortality rates.^22^ Thus, when it comes to improving health outcomes, using DHT to build trust in clinical trials could be particularly important for the Black US population.

In light of the potential benefits of utilizing DHTcd in clinical trials to enhance representativeness, it is also crucial not to overlook the significant challenges with regard to recruitment and retention of previously underrepresented patient groups. DHTs are more likely to be employed by patient groups that are already adequately represented in clinical trials.^18^ In the absence of effective measures to reduce the barriers for underrepresented patient groups to utilize DHT, there is a risk of a widening digital divide in clinical trials.^8,25,26^ In this regard, digital literacy, i.e., the knowledge and skills required to use DHT appropriately and in a risk- and opportunity-aware manner, plays a key role for successful recruitment and DHT uptake in respective clinical trials. Thereby, digital literacy is not randomly distributed across different patient groups: potential trial participants, e.g., in older age groups may be less likely to use DHT, as digital literacy is comparatively lower. In general, it can be assumed that some patient populations already underrepresented in conventional trials tend to have lower digital literacy and therefore exhibit even higher risks of being excluded from DHT-based trials.^27^

However, the challenges of using DHT in clinical trials are not limited to recruiting patient groups with limited access to DHT. To fully realize the benefits of DHT for clinical trials, high retention rates are also required. Even if representative samples are recruited at trial beginning, non-random attrition may mean that results cannot be generalized over a given trials’ course. Accordingly, available data demonstrate that in eight US DHT remote trials, more than half of the initial 100,000 participants dropped out within a week. Hence, dropout rates varied widely according to disease status, age, use of financial incentives, and clinical referral.^28^ As further findings from the Framingham Heart Study’s nested eCohort suggest, the likelihood of completing all DHT-based surveys of a particular trial period increases with higher levels of education and female gender.^29^ Apart from the latter two studies^28,29^, there is no empirical evidence on the suitability of DHTcd for enhancing clinical trial representativeness available. Using metadata from all clinical trials registered on clincaltrials.gov between 2013 and 2023, our study provides the lacking evidence:we evaluated whether female and Black participants as well as participants older than 65 years are more adequately represented in completed DHTcd trials than in non-DHT trials.

To assess whether differences in the evaluated participant groups already arise in access to DHTcd and non-DHT trials, we report differences in eligibility criteria and actual trial participant proportions. Furthermore, recording participant sample compositions is essential for adjusting the representativeness of trials if necessary. We therefore also report on group-specific differences in the reporting of sample compositions. Depending on its indication, the use of DHTcd in a trial may affect the representativeness of certain groups — such as women, Black participants, and adults over 65 — in different ways. The benefit of using DHTcd may therefore vary depending on specific therapeutic areas. Hence, we also report on group-specific differences in representations of DHTcd and non-DHT trials by therapeutic area.

## Results

### Characteristics and Demographic Reporting of Trials Included

US-based trials registered on ClinicalTrials.gov with trial status specified as ‘completed’ between January 2013 and December 2023 were included in the analyses. This corresponds to 68,206 out of 536,794 trials with a total of more than 3.4 billion reported participants registered on ClinicalTrials.gov until May 2025, as illustrated by Figure 1. Further, 7.42% of the analyzed trials utilized DHT in an unspecified manner. Following the classification as laid out in Marra et al.^30^, the use of DHTcd was observed in 66.79% of the trials classified as using DHT and in 4.96% of all clinical trials included.

**Figure 1.**
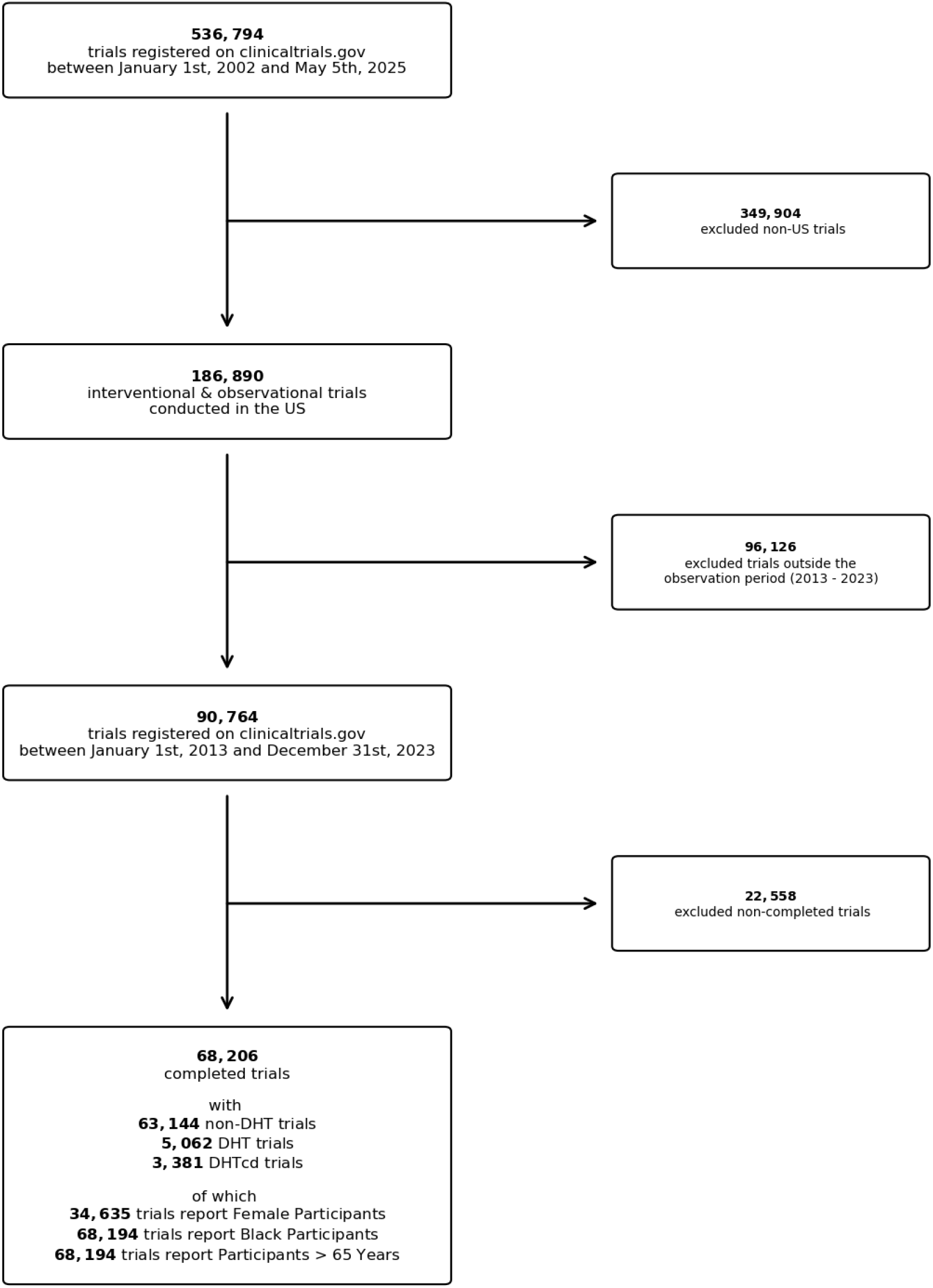
CONSORT Diagram of analyzed trials. Data on trials registered on ClinicalTrials.gov. was accessed through the aggregate analysis of ClinicalTrials.gov (AACT) database API.^31^

Information on shares of female participants was provided for 44.2% of the included trials. Also, information on shares of participants over 65 years of age was provided for 44.2% of the trials. The proportion of trials reporting both of these participant characteristics remained constant throughout the entire observation period (SD = 1.4% and SD = 1.4%). Following the introduction of the US final rule on January 18, 2017 that requires reporting of the proportion of trial participants by race and ethnicity,^32^ the proportion of trials stating the overall percentage of Black participants in trial populations increased from an average of 22.2% (SD = 4.6%) to 39.3% (SD 1.4%) per year. Overall, information on the shares of Black participants in trial populations was provided for 32.3% (SD = 9.7%) of the included clinical trials (see also Supplementary Figure A1).

As can furthermore be inferred from Figure 2, both proportions of DHT and DHTcd trials increased over the entire observation period, from 1.92% in 2013 to 7.46% in 2023 among DHTcd trials, and from 2.62% in 2013 to 11.22% in 2023 among DHT trials.

**Figure 2.**
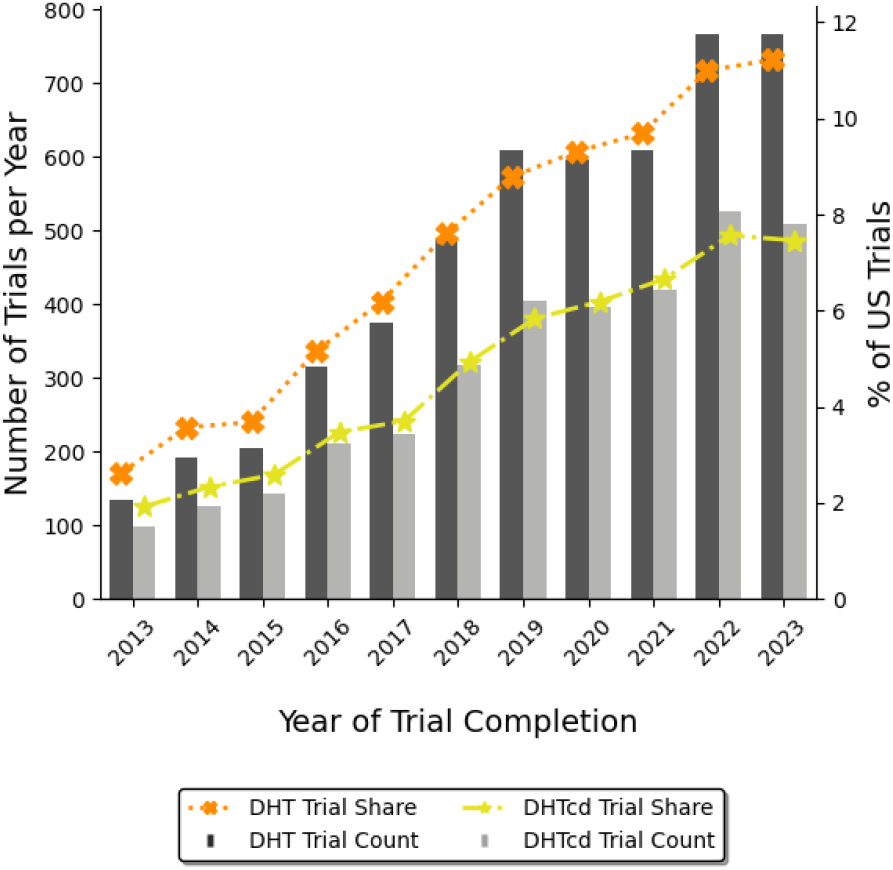
Total numbers and relative shares of DHT and DHTcd trials by year, 2013 to 2023. Relative shares of DHT and DHTcd trials were calculated in relation to all completed US trials registered in a given year. *Data source:* AACT.^31^

### Differences of Demographic Reporting between DHTcd and non-DHT Trials

With regard to reporting on shares of female and Black participants and shares of participants above the age of 65 years, differences were observed between DHTcd and non-DHT trials. Overall, 45.1 % (SD = 1.4%) of non-DHT trials reported shares of female participants, compared to 28.6% (SD = 3.5%) of DHTcd trials. As can be seen in Figure 3, the share of non-DHT trials reporting the share of female participants remained almost constant over the entire observation period. However, as also indicated by the larger standard deviation, there was larger variation in reporting on female participants in DHTcd trials.

**Figure 3.**
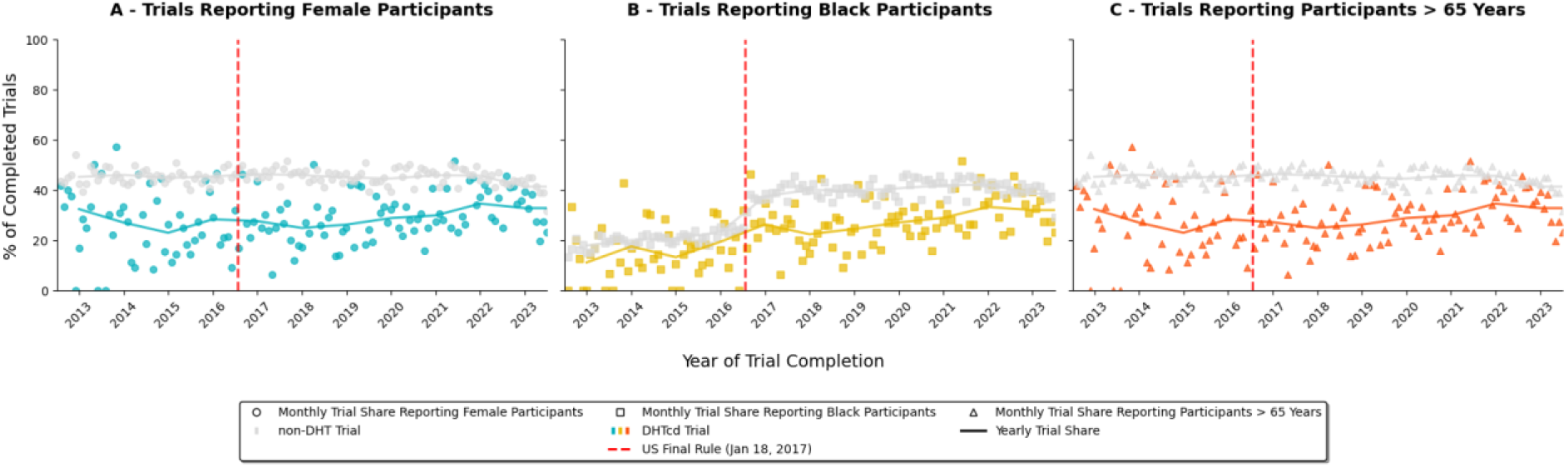
Completed DHTcd and non-DHT trials reporting demographic trial compositions, 2013 to 2023. **A** Trials reporting female participant shares. **B** Trials reporting Black participant shares. **C** Trials reporting participant shares older than 65 years. Circles, squares and triangles represent monthly shares of completed US trials reporting demographic participant sample compositions, while lines indicate yearly shares. Trial shares were calculated in relation to all completed US trials registered in a given month and year. The US Final Rule is indicated by the red dashed line. Effective since 18 January 2017, the US Final Rule issued by the FDA expands the reporting requirements for clinical trials on ClinicalTrials.gov to include race and ethnicity. *Data source:* AACT.^31^

A similar pattern emerges when examining reports of participant shares above the age of 65 years in trial populations. These participant shares are reported in 45.0% (SD = 1.4%) of the included non-DHT trials and in 28.6% (SD = 3.5%) of the DHTcd trials, with a more constant reporting for non-DHT trials compared to DHTcd trials.

We make different observations regarding the trials reporting Black participant shares of trial populations. Initially, a visual inspection of Figure 3 reveals an increase in non-DHT trials reporting the proportion of Black participants in trial populations from an average of 22.4% (SD = 4.8%) before the introduction of the final rule in January 2017 to an average of 40.0% (SD 1.7%) after its introduction. However, this increase is not as pronounced for DHTcd trials. The proportion of DHTcd trials with a higher proportion of Black participants increased from an average of 12.2% (SD = 7.6%) before introduction to an average of 27.7% (SD = 3.9%) after introduction. Overall, differences exist between DHT and DHTcd trials: the proportion of Black trial participants is reported in 32.9% (SD = 10.0%) of non-DHT trials as compared to 23.2% (SD = 7.3%) of the DHTcd trials.

### Differences of Inclusion Criteria between DHTcd and Conventional trials

Inclusion and exclusion criteria determine the access of population groups to clinical trials. Therefore, we analyzed the extent to which the shares of trials granting access to women and participants over 65 years differed between DHTcd and non-DHT trials. As a result, 96.78% of the DHTcd trials were designed to include female participants as part of their inclusion criteria, whereas trial access was granted to female populations by 96.32% of the non-DHT trials. Both proportions remained constant over time, as can be inferred from Figure 4. In 2013, 99.0% of the DHTcd were planned to include female populations, whereas inclusion criteria permitted female participants in 95.7% of the non-DHT trials in the same year. In 2023, 97.1% of the DHTcd trials admitted female participants as opposed to 96.5% in non-DHT trials. Overall, 34.46% of the DHTcd and 40.16% of the non-DHT trials were planned to include participants over 65 years old. As Figure 4 also illustrates, these proportions remained relatively constant over time. Accordingly, 32.3% of DHTcd trials and 37.6% of non-DHT trials in 2013 included participants older than 65 years of age. In 2023, participants above the age of 65 were included in 36.1% of the DHTcd trials and 42.1% of the non-DHT trials.

**Figure 4.**
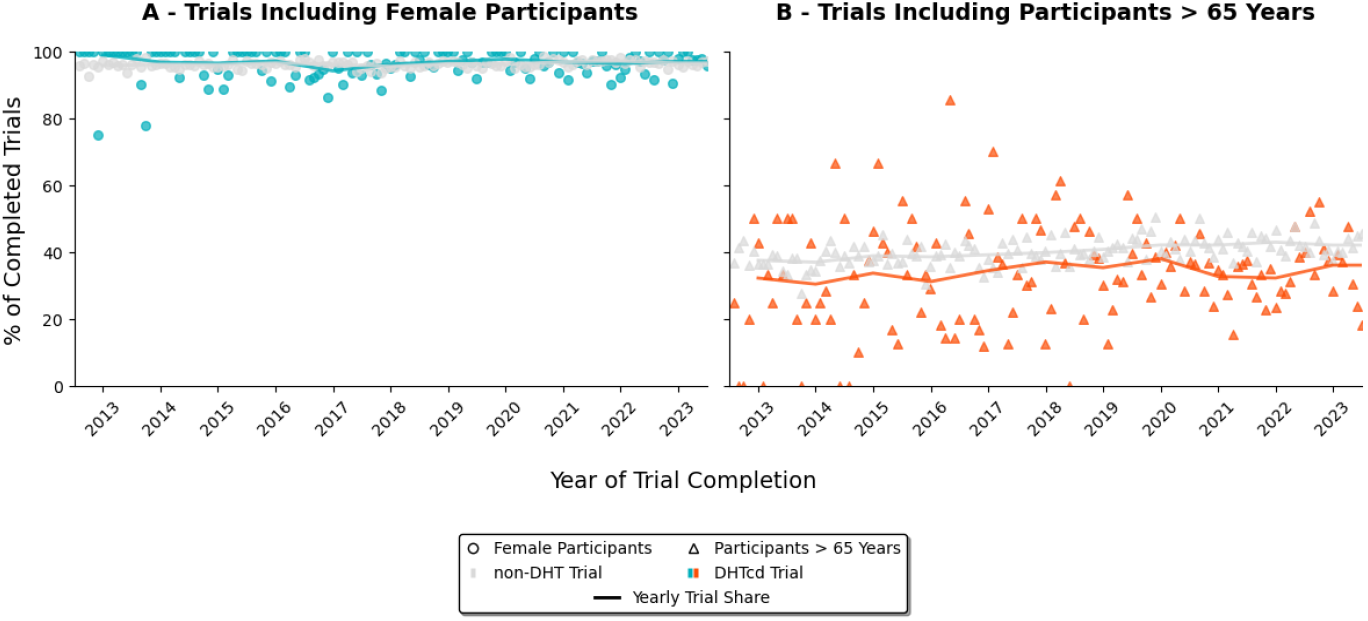
DHTcd and non-DHT trials including female participants and participants older than 65 years, 2013 to 2023. **A** Shares of trials including female participants. **B** Shares of trials including participants older than 65 years. Dots and triangles represent monthly shares of completed US trials, while lines indicate yearly shares. Trial shares were calculated in relation to all completed US trials registered in a given month and year. *Data source:* AACT.^31^

### Differences in Representation of DHT and Conventional Trials

Figure 5 shows differences in the composition of trial populations with regard to shares of female and Black participants and participants older than 65 years. The shares of these three groups within the US population are also shown and serve as a benchmark for determining trial representation. The proportion of women in the US population was 49.97% in 2013, whereas the median proportion of female participants in non-DHT studies was 43.75%. Female study participants were therefore underrepresented. With a median proportion of female participants of 45.00% in 2013, this also applies to DHTcd trials. In 2023, 49.75% of the US population was female. In contrast, the median proportion of female participants in all non-DHT trials was 47.32%. Hence, female participants in non-DHT trials were slightly underrepresented in 2023, too. However, this does not apply to DHTcd trials, for which a median proportion of 58.00% female participants indicates an overrepresentation in 2023.

**Figure 5.**
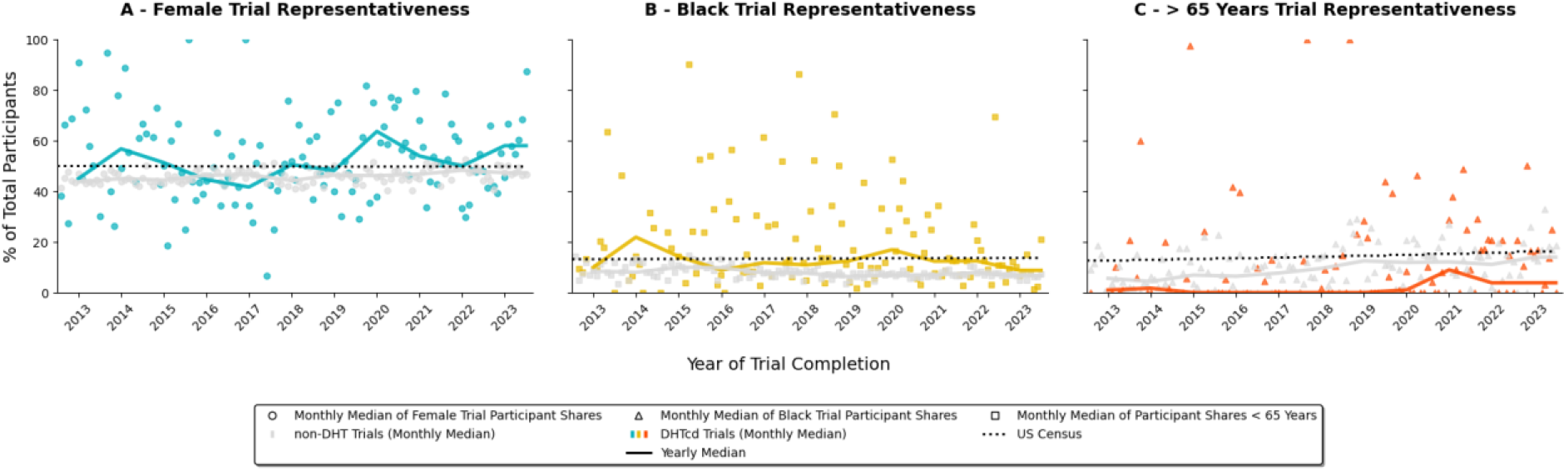
Demographic representation in DHTcd and non-DHT trials, 2013 to 2023. **A** Trial representativeness of female participants. **B** Trial representativeness of Black participants. **C** Trial representativeness of participants older than 65 years. Dots, squares and triangles represent monthly median shares of completed US trials, while lines indicate yearly median shares. Medians were calculated based on trials completed in a given month and year. US census shares were calculated in relation to the total US population in a given year indicated by the dotted line. *Data sources:* AACT^31^and US Census.^33^

Similar trends can be seen among Black trial participants. This group is also underrepresented in the medians of non-DHT studies, as measured by their US population share. Given Black US population shares of 13.18% in 2013 and of 13.66% in 2023, median Black participant shares of trial populations amounted to 8.44% and 6.67% in 2013 and 2023, respectively. Black trial participants are better represented in DHTcd, as median participant proportions of 10.00% in 2013 and of 8.79% in 2023 indicate.

However, a different picture emerges when looking at participants aged over 65 years. In light of US population shares of 12.63% in 2013 and 16.21% in 2023, this group is underrepresented in non-DHT trials throughout the observation period. In non-DHT trials, the median percentage of participants aged over 65 years was 5.59% in 2013 and 13.96% in 2023. Throughout the observation period however, yearly median proportions of participants over 65 years of age in DHTcd trials were consistently lower than in non-DHT trials: in 2013, the median proportion of participants in DHTcd trials aged over 65 years old was 1.02%, while we observed 3.86% in 2023 for the same group of participants.

Trial representativeness gaps were calculated in order to obtain information on whether DHTcd is associated with better representativeness of the three analysed groups of female and Black study participants and study participants over 65 years of age. Accordingly, panel A of Figure 6 shows that the use of DHTcd in female study participants leads to a median overrepresentation of 1.7%. This contrasts with an underrepresentation of female study participants of -3.8% in non-DHT trials. There was also a better median representation of Black study participants in trials in which DHTcd was used (−1.6% in DHTcd vs. -5.8% in non-DHT trials). However, the same applies to study participants over the age of 65: in DHTcd trials, the group of participants older than 65 years is underrepresented at -14.2%. This contrasts with an underrepresentation of -5.4% in non-DHT trials.

**Figure 6.**
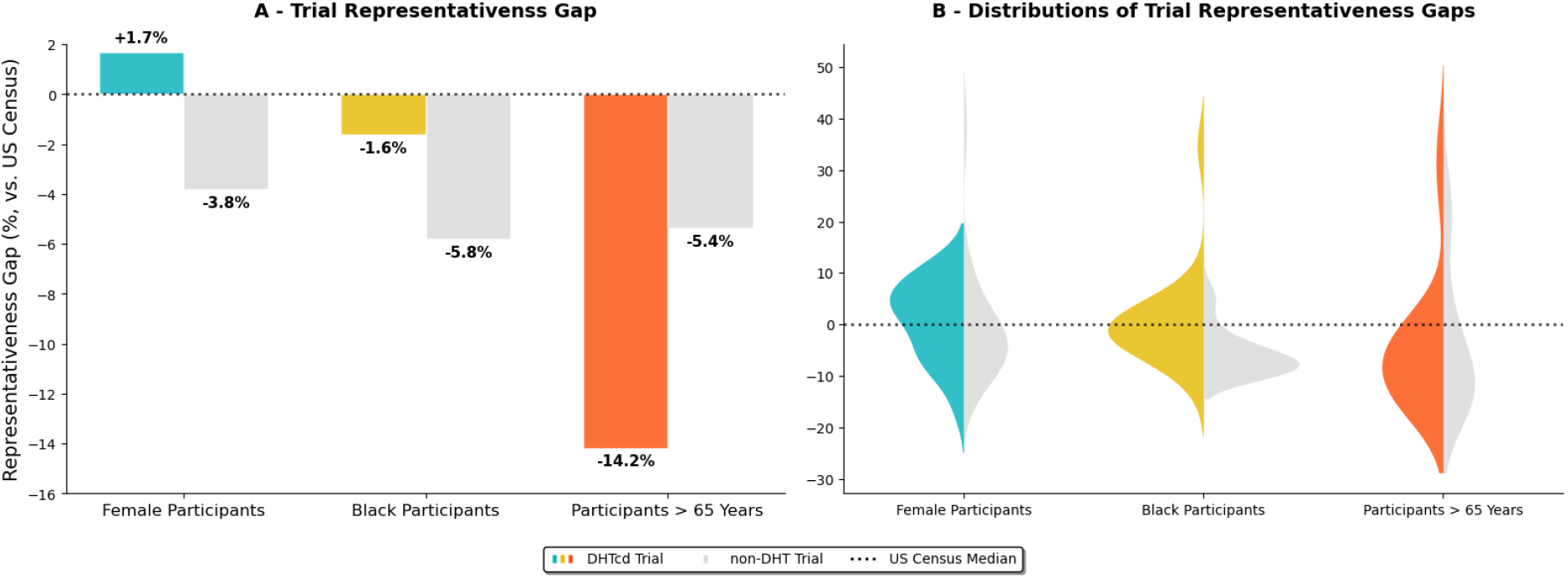
Representation gaps in DHTcd and non-DHT trials. **A** Trial representativeness gaps are shown as differences between median trial participant shares and corresponding US census medians by demographic category and trial type. **B** Distributions of representation gaps by demographic category and trial type. Median trial participant shares were calculated based on all US trials completed between 2013 and 2023. As indicated by the black dotted line, US census medians were calculated based on yearly census shares from 2013 to 2023. *Data sources:* AACT^31^ and US Census.^33^

Panel B of Figure 6 depicts the distribution of representation gaps for each of the three analysed participant groups. Of these three participant groups, female trial participants exhibit the most consistent distribution pattern. In both DHTcd and non-DHT trials, this group shows the fewest extremes in representation gaps. Among Black participants, more trials exhibit extreme representation gaps in DHTcd than in non-DHT trials. Compared to the other two groups, a higher number of DHTcd and non-DHT trials with extreme representation gaps is found in the group of participants aged over 65.

### Trial Representativeness Improvements by Therapeutic Area

Compared to DHT trials, the median deviation in the proportion of female participants in DHTcd trials is highest in the following three therapeutic areas, as Figure 7 indicates: ‘Diseases of the genitourinary system’ (+27.9%), ‘Diseases of the musculoskeletal system or connective tissue’ (+8.7%) and ‘Mental, behavioural or neurodevelopmental disorders’ (+6.3%). As can further be seen in Figure 7, DHTcd trials are associated with higher representation than DHT trials in 9 out of 21 analysed therapeutic areas for female participants. DHTcd trials in the remaining 12 therapeutic areas showed lower representativeness than non-DHT trials.

**Figure 7.**
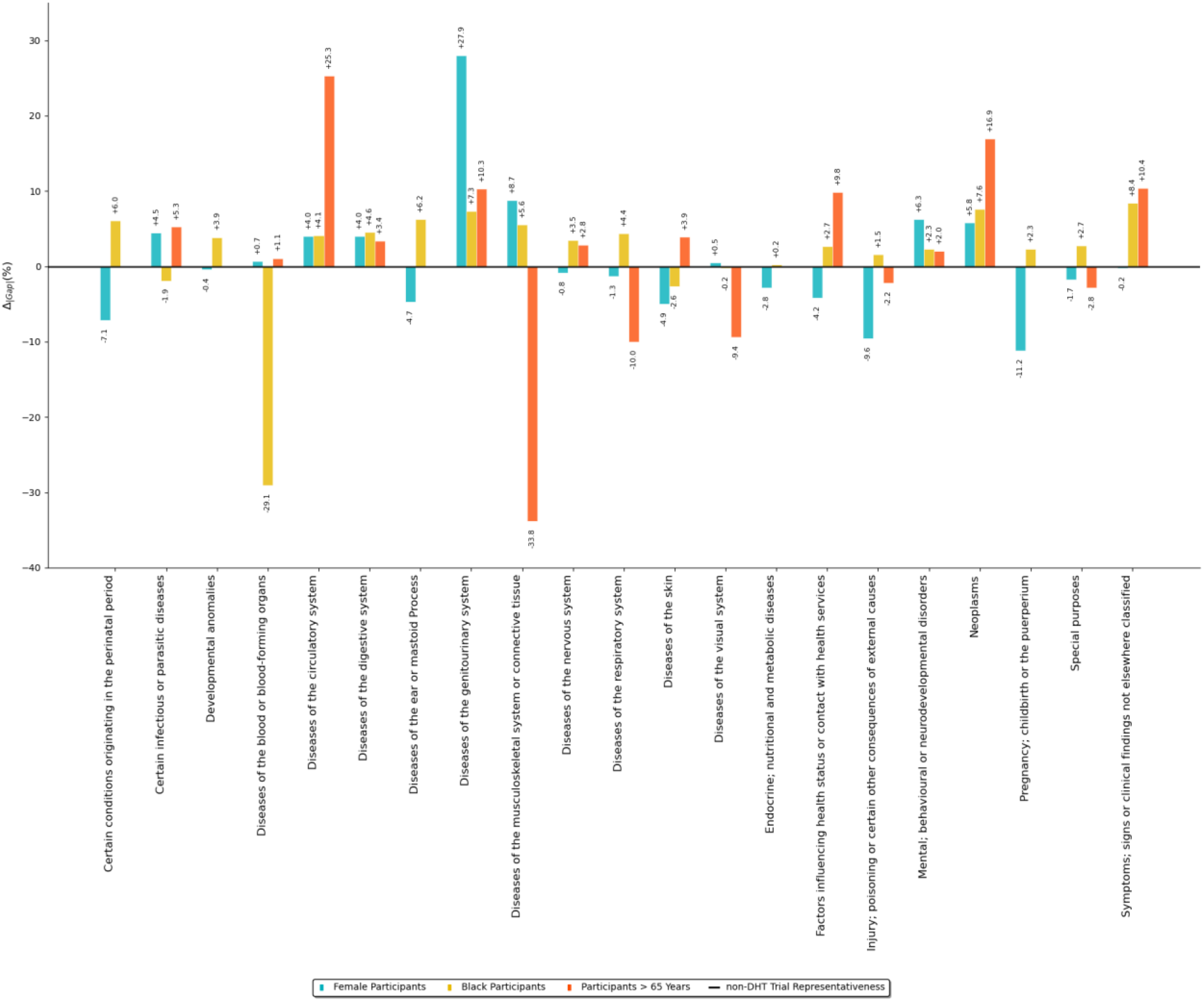
DHTcd vs. non-DHT Trial Representativeness Gaps by Therapeutic Area. Differences in absolute representation gaps were calculated as Δ_|**Gap**|_= |median(non-DHT trial participant shares) ‐ median(US Census share)| ‐ |median(DHTcd trial participant shares) ‐ median(US Census share)| per therapeutic area. Median trial participant shares were calculated based on all US trials completed between 2013 and 2023. US census median shares were calculated based on yearly census shares from 2013 to 2023. Positive values indicate by how many percentage points median shares of participants in DHTcd trials deviate less from the corresponding US Census median than in non-DHT trials. Negative values indicate by how many percentage points DHTcd trials deviate more from the US census median than non-DHT trials. *Data sources:* AACT^31^ and US Census.^33^

In 4 out of 21 therapeutic areas, non-DHT trials exhibit a higher share of Black participants. DHTcd trials show a higher proportion of Black participants in 17 of the 21 therapeutic areas (Figure 7). In the following three therapeutic areas, we observe the highest median deviation in the shares of Black participants in DHTcd trials compared to non-DHT trials: ‘Symptoms, signs, or clinical findings not elsewhere classified’ (+8.3%), ‘Neoplasms’ (+7.6%), and ‘Diseases of the genitourinary system’ (+7.3%).

In most cases, median shares of participants aged over 65 years in DHTcd trials deviate less from the corresponding US Census shares. Accordingly, the trials with the highest representativeness of participants older than 65 years are represented in the ‘Diseases of the circulatory system’ (+25.3%), ‘Neoplasms’ (+16.9%), and ‘Symptoms; signs or clinical findings not elsewhere classified’ (+10.4%) therapeutic areas. As also shown in Figure 7, median deviations from the US Census of DHTcd trials participant shares are higher as compared to non-DHT trials in six therapeutic areas.

## Discussion

Our study demonstrates the potential of DHTcd to improve clinical trial representativeness. The potential arises from the possibility of using DHT to collect data with minimal technical barriers, continuously and regardless of participant location. In this context, it can be assumed that the use of DHTcd not only facilitates the implementation of decentralised trials, but also helps to strengthen willingness to participate in clinical trials as it fosters trust in healthcare institutions.^3,9^ Because of these characteristics, the use of DHTcd may be particularly beneficial for previously not adequately represented populations to expand population-specific biomedical knowledge. However, this reasoning is in contrast to the findings that group-specific sample compositions are recorded and reported less frequently in DHTcd trials than in non-DHT trials. These findings rather suggest that DHTcd - despite increasing use - are not primarily used to monitor and possibly improve representation of the three investigated population groups in trial samples.

Nevertheless, in line with the assumption that using DHTcd increases confidence and willingness to participate in clinical trials, the present results show that DHTcd trials are associated with higher representativeness than non-DHT in all of the three analysed participant groups for the most relevant therapeutic areas, as measured by group-specific years of life lost (YLL): 9 therapeutic areas in which using DHTcd is associated with higher representativeness cover 10 out of 21 Global Burden of Disease (GBD) Level 2 risk factors of YLL within the female participant group. These 21 risk factors also correspond to 12 therapeutic areas where DHTcd trials are associated with higher representation of the participant group older than 65 years.^34^ Similarly, 10 therapeutic areas in which the use of DHTcd leads to better representation of Black trial participants correspond to the 10 highest YLL risk factors within this US population group.^35^

These results are contrasted by the finding that, when considered as a whole, the group of trial participants older than 65 years is less represented when data is collected using DHTcd. A possible explanation for this finding is offered by the fact that deviations from adequate representations tend consistently more towards the extremes in DHTcd trials as compared to non-DHT trials. In light of such larger extremes in representation gaps, a potential reason for the comparatively lower representation of DHTcd trial participants over 65 years of age is a digital divide.^25^ Such a digital divide may possibly result from differences in DHTcd utilization between the investigated participant groups. Older people are less likely to be digitally literate and willing to use DHT.^36,37^ This fact possibly also explains why people over the age of 65 have so far been included less frequently in DHTcd trials than in non-DHT trials. Assuming that this fact is known or anticipated by sponsors and research organisations, this also explains our finding that people over the age of 65 have been included less frequently in DHTcd studies than in non-DHT studies.

The greater variability in the representation of DHTcd trial participants suggests that the use of DHTcd cannot be expected to generally enhance trial representativeness, despite its potential. As regulatory agencies increasingly emphasize diversity in clinical development programs, sponsors must carefully consider how digital trial designs might inadvertently exclude patient populations. Without proactive measures to address digital literacy gaps and ensure equitable access to technology, the potential of DHT to enhance inclusion in clinical research may instead exacerbate existing disparities. Therefore, implementing DHTcd in clinical trials requires assessments of the factors influencing usage behaviour to ensure reliable participant representation. In addition to the compound effects of user characteristics, specific trial settings and DHTcd properties must be considered as part of such assessments. Research in this domain must also acknowledge that distinct therapeutic areas may necessitate tailored DHTcd approaches, whether deployed individually or in combination. This variability inherently creates divergent demands upon study participants, whose capacity to meet these challenges is contingent upon factors including digital literacy, trust, and underlying disease status. Findings from such research are fundamental to guiding future developments of DHTcd trials. These evidence-informed trial designs will facilitate robust estimation of subgroup-specific treatment efficacies while advancing the development of therapeutic interventions that demonstrate both safety and effectiveness across all patient populations.

## Methods

### Identification of DHT and DHTcd trials

Our descriptive analyses are based on data from a total of 536,794 clinical trials between January 1^st^, 2002 and May 5^th^, 2025 for which metadata was available on ClinicalTrials.gov. The trial metadata was downloaded on 05/09/2025 using SQL queries via the aggregate analysis of ClinicalTrials.gov (AACT) database API.^31^ Trials utilizing DHT were flagged as according to Marra et al.^30^ and Marra & Stern^38^ by searching ClinicalTrials.gov’s title, name, intervention, description, and measure fields for DHT product names. DHT products were identified after updating the lists used in Marra et al.^30^ and Marra & Stern^38^ with a curated list of DHT issued by ICON PLC as^1^ of February 18th, 2025. To minimise the rate of false positive flags, the updated list was visually inspected and product names that were ambiguous in medical terminology were replaced with the name of the product manufacturer. For example, ‘ACE’ as the name for Fitbit’s ACE smartwatch was replaced with ‘Fitbit’ to prevent conventional trials evaluating ACE inhibitors from being flagged as DHT trials. The updated list included 4,183 names of DHT products or manufacturers. According to Marra et al.^30^, the use of a DHT in any given clinical trial can be classified as one of four mutually exclusive use cases. Per this classification, only one of these four use cases involves the use of DHTs not as an intervention, but to capture endpoint data for another intervention or data of interest in an observational study. DHT trials corresponding to this use case (e.g. DHTcd trials) were therefore flagged as trials in which the intervention type was not a device, and no match was found between the intervention name and a DHT product name.

### Sociodemographic Category Mapping

ClinicalTrials.gov provides customizable categories and category titles for mutually exclusive and exhaustive information on sociodemographic characteristics of trial participants. Sociodemographic characteristics are reported as participant counts. To identify information on age, sex, and race/ethnicity, clinicaltrial.gov’s category title field was first fuzzy searched for ‘Race’, ‘Ethnicity’, ‘Sex’, ‘Gender’, ‘Male’, ‘Female’, ‘Age’ keywords. Items in the resulting list were assigned to the new category titles ‘Age’, ‘Sex’, ‘Race/Ethnicity’ after visual inspection.

Race and ethnicity categories were then merged to match the categories (as classified by the US Census Bureau) most commonly reported in trials: White, Hispanic, Black, Asian (including Pacific Islanders and Native Hawaiians), and American Indian (including Alaska Natives).^39^ This resulted in a total of 17,786 different categories that were mapped onto the 17 distinct categories used by the National Institutes of Health (NIH) for age, sex, and race/ethnicity data. LLaMa 3.3 - 70B - Instruct was used for mapping. To assess the mapping quality, a random sample of 100 categories was annotated independently by two annotators for each of the ‘Age’, ‘Gender’ and ‘Race/Ethnicity’ categories. Based on the annotated categories lists, LLaMa 3 - 8B – Instruct (F1 score 0.76) as well as openAI’s GPT-4o-mini (F1 score 0.83) and GPT-4o (F1 score 0.85) showed inferior performance when compared to LLaMa 3.3 - 70B - Instruct (F1 score 0.90). See also Table T1 in the appendix for details.

The analyses of trial representation in terms of race/ethnicity categories followed the same approach as in Turner et al.^39^ In most studies, Hispanic identity is recorded as a separate variable from race, less frequently in combination with race (e.g. ‘White, Hispanic’). In the latter case, persons recorded as ‘White’ cannot be distinguished from persons recorded as ‘Latino’. Such trials were excluded from the analyses.

### Therapeutic Area Mapping

Therapeutic areas were first mapped to the Medical Subject Headings (MeSH) entries in the AACT database, and then to the Concept Unique Identifiers (CUIs) and finally MeSH IDs included in the Unified Medical Language System (UMLS) Metathesaurus, using the UMLS Terminology Services Crosswalk REST API. The CUI was then used to assign the appropriate ICD-10 codes via the UMLS. These mappings were supplemented with ICD-10 mappings from the Disease Ontology (DO) project of the International Health Terminology Standards Development Organisation (IHTSDO) via the assigned MESH IDs, in order to increase the proportion of trials with assigned ICD-10 codes.^40^ ICD-10 codes were then used to determine therapeutic areas, ensuring comparability of ICD code assignments over the entire observation period from 2013 to 2023.^41^ Designations for therapeutic areas were created based on the ICD-10 chapter definitions.

## Data Availability

All data produced in the present study are available upon reasonable request to the authors.

## Competing Interests

The authors declare no competing interests.

## Author Contributions

AFN conceptualized the study, wrote the code for data preprocessing and analyses and wrote the manuscript. LS conceptualized the study, contributed code for data processing and supported the conceptualization of the study. MK contributed code for data preprocessing and critically reviewed the manuscript. MSA contributed code for data preprocessing. AA conceptualized and contributed code for data preprocessing and analyses.

## Appendix

**Table T1:**
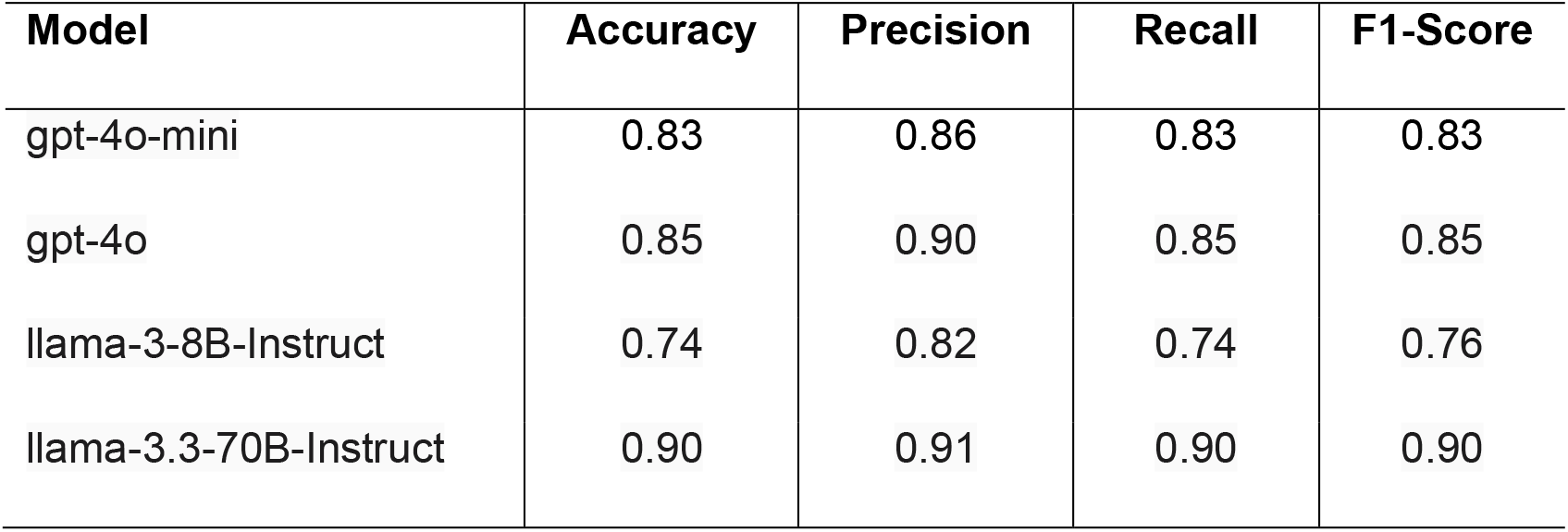
Performance metrics of LLM’s used for mapping of age, gender and ethnicity categories. Data Source: AACT.^31^

| Model | Accuracy | Precision | Recall | F1-Score |
| --- | --- | --- | --- | --- |
| gpt-4o-mini | 0.83 | 0.86 | 0.83 | 0.83 |
| gpt-4o | 0.85 | 0.90 | 0.85 | 0.85 |
| llama-3-8B-Instruct | 0.74 | 0.82 | 0.74 | 0.76 |
| llama-3.3-70B-Instruct | 0.90 | 0.91 | 0.90 | 0.90 |

**Figure A1:**
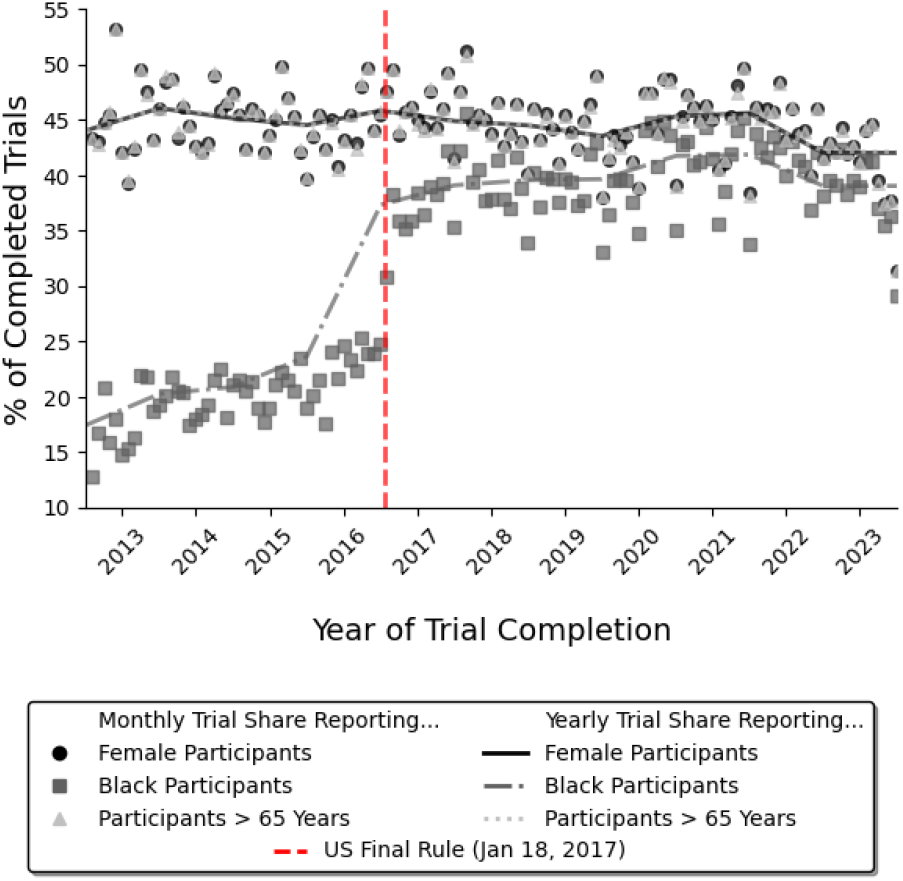
Completed US trials reporting demographic trial compositions, 2013 to 2023. Circles, squares and triangles represent monthly shares of completed US trials reporting, while lines indicate yearly shares. Trial shares were calculated in relation to all completed US trials registered in a given month and year. The US Final Rule is indicated by the red dashed line. Effective since 18 January 2017, the US Final Rule issued by the FDA expands the reporting requirements for clinical trials on ClinicalTrials.gov to include race and ethnicity. *Data source:* AACT.^31^

**Figure A2:**
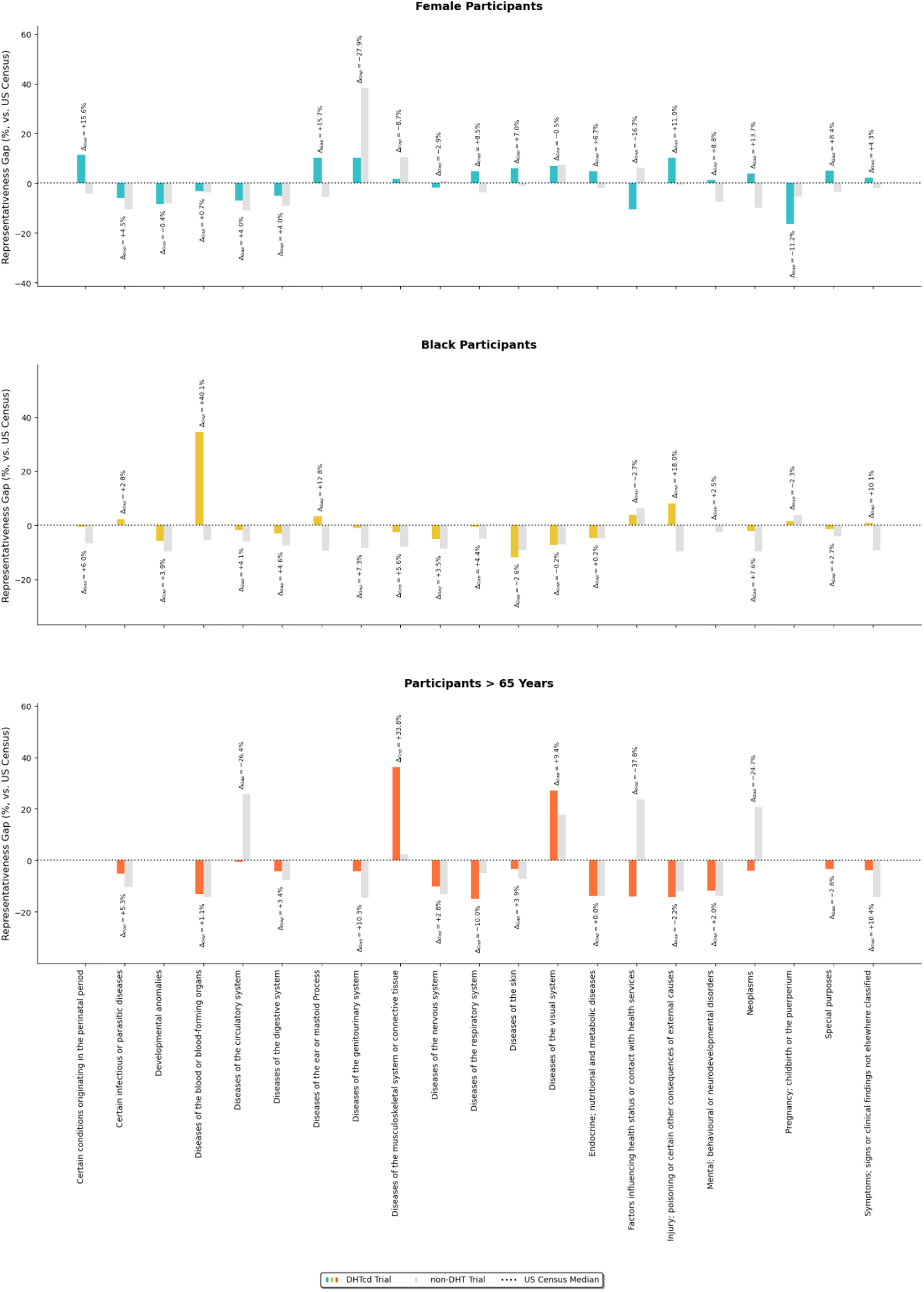
Representation gaps of DHTcd and non-DHT by Therapeutic Area. **a** Difference in representation gaps for female participants. **b** Differences in representation gaps for Black participants. **c** Differences in representation gaps for participant over the age of 65 years. Signed differences between representation gaps were calculated as Δ_**Gap**_ = (median(DHTcd trial participant shares) – median(US Census share)) – (median(non-DHT trial participant shares) – median(US Census Share)) per therapeutic area. Median trial participant shares were calculated based on all US trials completed between 2013 and 2023. US census median shares were calculated based on yearly census shares from 2013 to 2023. *Data sources:* AACT^31^ and US Census.^33^

## Footnotes

1 **ICON is a global provider of development and commercialization services for drugs and devices; we used their expertise to obtain a curated list of digital health technologies:** https://www.iconplc.com/solutions/outcome-measures/digital-health-technologies

## Notes

### Competing Interest Statement

The authors have declared no competing interest.

### Funding Statement

This study did not receive any funding.

